# Multi-trait association studies discover pleiotropic loci between Alzheimer’s disease and cardiometabolic traits

**DOI:** 10.1101/2020.08.26.20179366

**Authors:** William P. Bone, Katherine M. Siewert, Anupama Jha, Derek Klarin, Scott M. Damrauer, the VA Million Veteran Project, Kyong-Mi Chang, Philip S. Tsao, Themistocles L. Assimes, Marylyn D. Ritchie, Benjamin F. Voight

## Abstract

Identification of genetic risk factors that are shared between Alzheimer’s disease (AD) and other traits, i.e., pleiotropy, can help improve our understanding of the etiology of AD and potentially detect new therapeutic targets. Motivated by previous epidemiological correlations observed between cardiometabolic traits and AD, we performed a set of bivariate genome-wide association studies coupled with colocalization analysis to identify loci that are shared between AD and eleven cardiometabolic traits. We identified three previously unreported pleiotropic trait associations at known AD loci as well as four novel pleiotropic loci. One associated locus was tagged by a low-frequency coding variant in the gene *DOCK4* and is potentially implicated in its alternative splicing. Statistical colocalization with expression quantitative trait loci identified by the Genotype-Tissue Expression (GTEx) project identified additional candidate genes, including *ACE*, the target of the hypertensive drug class of ACE-inhibitors. We found that the allele associated with decreased *ACE* expression in brain tissue was also associated with increased risk of AD, providing human genetic evidence of a potential increase in AD risk from use of an established anti-hypertensive therapeutic. Overall, our results support a complex genetic relationship between AD and these cardiometabolic traits, and the candidate causal genes identified suggest that blood pressure and immune response play a role in the pleiotropy between these traits.

## Introduction

Studies have consistently found a positive epidemiological correlation between Alzheimer’s disease (AD) and cardiometabolic traits, yet the biological mechanisms behind this correlation is not well understood^1-4^. A leading hypothesis is that this correlation is due to shared genetic influence, or pleiotropy, between AD and cardiometabolic traits^4^. By identifying pleiotropic loci between these traits, we can (i) identify new therapeutic targets or opportunities for drug repurposing, (ii) predict potential side effects, and (iii) better understand the etiology of these complex traits. The identification of new therapeutic targets for AD is of particular importance since AD afflicts approximately 50 million people, and there exist only a handful of therapeutics available for AD that have only limited efficacy in slowing the progression of the disease^5^.

Pleiotropy has been an area of both theoretical and empirical study at least since the beginning of the twentieth century^6-8^. However, the topic has received renewed attention, given the pervasiveness of pleiotropy that has been uncovered through genome-wide association studies (GWAS)^8-11^. Recent methods and analysis have sought to characterize the extent of the phenomenon throughout the genome^8,12-15^, quantifying pairwise genetic correlation across a battery of traits^8,10,16^, exploiting pleiotropy to perform causal inference in the framework of Mendelian randomization^8,17-19^, or statistically co-localizing association signals across two or more traits^8,20-22^. These methods and publicly available GWAS summary statistics enable studies to dissect the shared genetic etiology between AD and cardiometabolic traits^8,12^. Due to the epidemiological correlation between AD and cardiometabolic traits, coupled with the fact that many cardiometabolic traits are genetically correlated with one another, additional broader-scale pleiotropic studies are warranted, and recently the field has begun to do so^4,16^. Statistical methods for detecting pleiotropy use the definition of a single locus associated with two or more traits, and these methods are generally intended to detect loci that have a single genetic variant underlying the shared heritability at the locus.

However, recent studies have shown that at some pleiotropic loci there is no shared causal SNP, but instead different SNPs are causal for the different traits. These loci are associated with multiple traits but there is no shared causal genetic variant behind the associations^9,23^. For this reason, we consider here a more stringent definition of pleiotropy: loci that are associated with two or more traits, *and* where the statistical data provides evidence of a shared causal genetic variant. We used colocalization analysis to identify which loci appear to share causal genetic variants and which appear to be cases of spurious pleiotropy^8,20,21^.There are two models of pleiotropy for this scenario^8^. The first is horizontal pleiotropy, where a genetic variant has a direct effect on two or more traits. The other is vertical pleiotropy, where a genetic variant has a direct effect on a trait and a mediated effect on a second trait through the first trait^8^.

In this study, we used summary statistics from the largest publicly available single-trait GWAS to investigate pleiotropy between AD and eleven cardiometabolic traits using the metaMANOVA bivariate GWAS method followed by colocalization analysis^12,24,25^. This bivariate GWAS method takes summary statistics for two traits as input and performs a GWAS for the pair of traits, while taking the correlation across association statistics into account^12,24,25^. We used this method to perform two different experiments. The first experiment was an “AD-centric” analysis, intended to detect loci that are associated with AD, but previously not shown to be pleiotropic for cardiometabolic traits. We also performed a locus discovery analysis to discover loci that are not previously reported to be associated with either AD or the cardiometabolic trait.

### Methods

We performed two bivariate GWAS experiments intended to detect loci that are pleiotropic between AD and cardiometabolic traits. For ease of reproducibility, we first performed a pairwise bivariate GWAS between AD and each of eleven cardiometabolic traits for both experiments. We then assessed whether there was evidence of a shared causal SNP at each bivariate significant locus by performing a colocalization analysis between the AD and cardiometabolic trait signals. To identify candidate causal genes, we performed colocalization analyses between the pleiotropic signals and single-tissue eQTLs from Genotype-Tissue Expression (GTEx) project v7^22^.

### Bivariate GWAS

We used the summary statistics from publicly available single-trait GWAS to performed pairwise metaMANOVA bivariate GWAS between AD^26^ and the following cardiometabolic traits: coronary heart disease (CHD)^27^, type II diabetes (T2D)^28^, systolic blood pressure (SBP)^29^, diastolic blood pressure (DBP)^29^, body mass index (BMI)^30^, waist-hip ratio adjusted for BMI (WHRadjBMI)^31^, body fat percentage (BFP)^32^, total cholesterol (TC)^33^, low-density lipoproteins (LDL)^33^, high-density lipoproteins (HDL)^33^, and triglycerides (TG)^33^ (Table S1 and Table S2, Data Availability). For each of these studies, approval by an institutional review committee was obtained, and all subjects gave informed consent, as documented in each original publication. All bivariate GWAS were performed using the *bivariate_scan* software^12,24,25^. Each bivariate GWAS resulted in a set of independent loci, which we defined as the genomic region that includes all SNPs within 1 MB of the bivariate lead SNP and any other SNPs that are in LD of r^2^ > 0.2 with the lead SNP using the 1000 Genomes European ancestry cohort (1kG EUR)^34^. Further detail on our bivariate GWAS pipeline can be found in the Supplemental Materials and Methods ^12^,^24^,^25^.

### AD-centric Analysis

We performed an AD-centric analysis to identify loci that are known to be associated with AD, but not previously known to be pleiotropic for cardiometabolic traits. We first performed pairwise bivariate GWAS between AD and each cardiometabolic trait (Table S3). To reduce the list of bivariate GWAS genome-wide significant loci results to just the loci that are near genome-wide significantly associated with AD and potentially associated with a cardiometabolic trait, we applied a filter that required loci to have an AD P-value < 1×10^-6^ and a cardiometabolic trait P-value < 5×10^-3^ (Figure 1).

**Figure 1.**
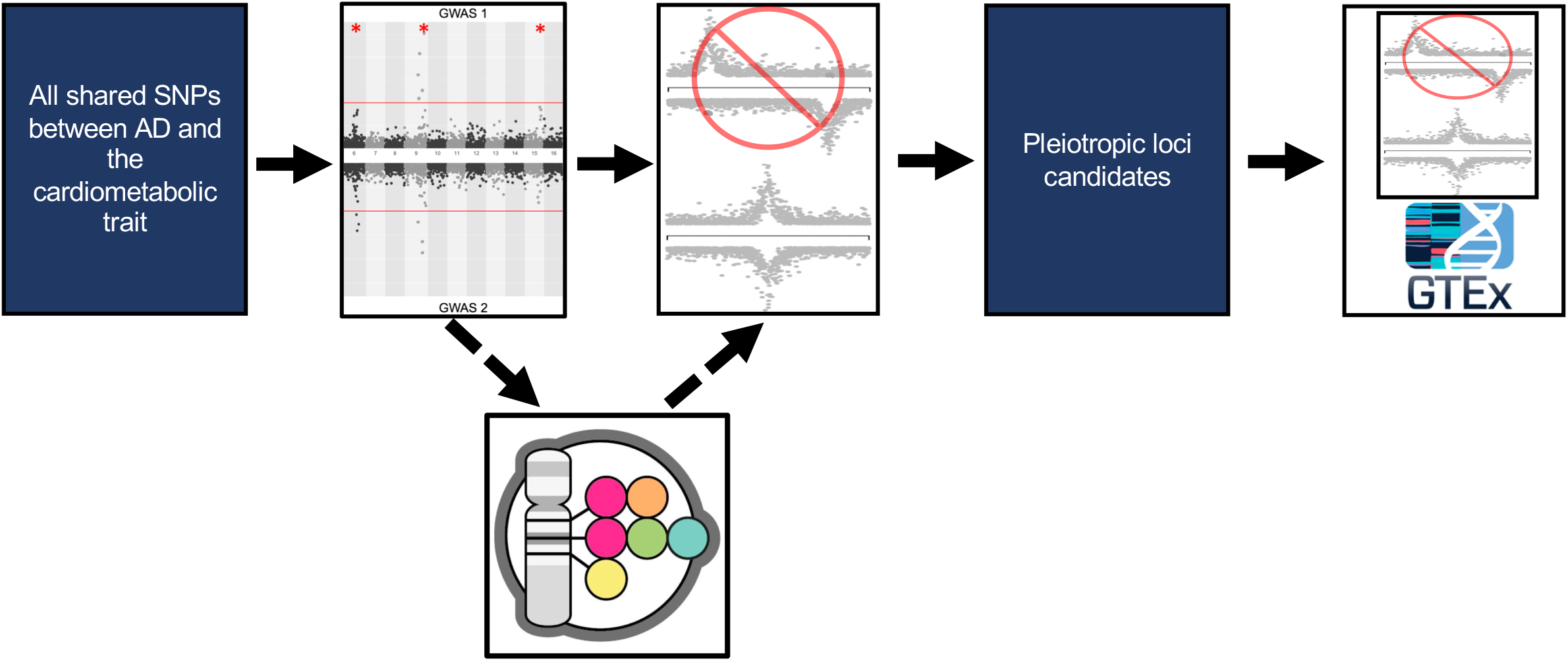
AD and cardiometabolic trait bivariate GWAS workflow Starting with all the SNPs that were in both GWAS summary statistics files, we performed a bivariate GWAS and filtered the bivariate significant loci based on their single trait P-values. For the locus discovery experiment, we removed loci that were in LD (1kG EUR r^2^ > 0.2) or within 500 KB of a known AD or the cardiometabolic trait being tested according to the GWAS Catalog (dotted line arrows). The filtration steps were followed by trait-trait colocalization to confirm there was evidence of a shared causal SNP between the signals at each locus. Finally, we performed single-tissue-eQTL analysis to identify candidate causal genes for each locus.

### Locus Discovery Analysis

To performed a locus discovery analysis we performed a bivariate GWAS between AD and each cardiometabolic trait (Table S4).To identify loci that were both pleiotropic and novel, we required the bivariate GWAS lead SNP had r^2^ < 0.2 in 1kG EUR and was greater than 500 KB away from all known single-trait associated loci for AD or the cardiometabolic trait being tested, as well as any loci from previous pleiotropic GWAS between the two traits^4,35^. Additionally, each locus needed to have at least a nominal single trait association with both traits, so we required an AD P-value < 5×10^-3^ and a cardiometabolic trait P-value < 5×10^-3^ (Figure 1).

### Trait-trait Colocalization

We performed colocalization analysis between the AD and the cardiometabolic trait signals given a 500 KB window (+/- 250 KB) around each locus using *COLOC*^20^. Our threshold for this analysis was a conditional probability of colocalization (i.e., PP4/ (PP3 + PP4) >= 0.8), which is defined as the posterior probability of colocalization conditioned on the presence of a signal for each trait (Figure 1). Loci that had a conditional probability of colocalization > 0.45 and < 0.8 were visually inspected using *LocusZoom* plots, and if the LD structure suggested additional associations unlinked to the leading variant in the region, we performed approximate conditional analysis (see Materials and Methods:

Approximate Conditional Analysis, below)^36^. We excluded loci in the *HLA* region and near the *APOE* locus from these experiments due to the difficulty in interpreting the independent contribution of these loci to these traits.

### Single-Tissue-eQTL Colocalization

We performed single-tissue eQTL colocalization analysis to prioritize candidate causal genes implicated by the pleiotropic signals detected in our bivariate GWAS. We collected the list of genes and tissues for which each bivariate GWAS lead SNP was a significant single-tissue eQTL in GTEx v7 from the GTExPortal (Tables S5-S10) (data from GTEx as of 02-28-2018, v7)^40^. We then performed colocalization using the AD association data at each locus and each single-tissue eQTL signal from GTEx v7 using a 500 KB window (+/- 250 KB) around the lead SNP using *COLOC^20^* (Figure 1). As above, we considered the AD and eQTL signals to colocalize if the conditional probability of colocalization was >= 0.8. We visually inspected the loci where the colocalization analysis resulted in a standard probability of colocalization < 0.8, but conditional probability of colocalization met our criteria^36^. For these loci we performed approximate conditional analysis, when the LD structure suggested there could be allelic series (see Methods: Approximate Conditional Analysis, below).

### Approximate Conditional Analysis

At each locus, we performed approximate conditional analysis on SNPs that appeared to be associated with the trait of interest independently of the lead SNP, because the presence of multiple associated variants in a region violates the assumptions of *COLOC^20^*. We identified potential nearby association signals using *LocusZoom* plots and the *LDassoc* tool of *LDlink*^36,37^. For each locus, we performed approximate conditional analysis using *GCTA-COJO* with 1000 Genome Project data (European samples, n=503) as a reference panel^38,39^. We conditioned our lead SNP on the most associated SNP for each potential confounding signals we identified at the locus. We then repeated the colocalization experiment on the locus using the conditional SNP P-values. We provide a full list of traits and loci we performed conditional analysis on, the lead SNP for each analysis, and the SNPs we conditioned on for each analysis are in the supplement (Table S11).

## Results

### AD-centric Analysis Results

We performed an AD-centric analysis to detect known AD loci that were not previously known to be pleiotropic with eleven cardiometabolic traits (Materials and Methods). We identified a total of 39 independent loci that were bivariate genome-wide significant, met our AD-centric single-trait P-value threshold of P-value < 1×10^-6^ and a cardiometabolic trait P-value < 5×10^-3^, and were outside of the *HLA* and *APOE* regions (Table S3).

We next performed trait-trait colocalization analysis on all 39 bivariate genome-wide significant loci to identify the subset of loci with evidence of a causal SNP shared in common between the AD signal and the cardiometabolic trait signal. Three loci met our colocalization criteria (Table 1). All of these loci are novel pleiotropic loci between AD and the respective cardiometabolic traits, but have previously been identified as genome-wide significant for AD in recent single-trait AD GWAS^4,26,41^.

**Table 1.** AD-centric analysis pleiotropic loci Chr, chromosome of the SNP; Direction of effect first position is the direction of effect of the effect allele on AD and the cardiometabolic trait; Effect Allele Frequency from the Jansen et al. 2019 allele frequency; Conditional Posterior Probability of Colocalization, PP4/ (PP3 + PP4) the results of the trait-trait colocalization analysis.

| Cardiom<br>etabolic<br>Trait | Locus<br>Name | Lead SNP | Chr | Position<br>GRCh37 | Effect<br>Allele/<br>Other<br>Allele | Directi<br>on of<br>Effect<br>AD/CM | Effect<br>Allele<br>Freque<br>ncy | Bivari<br>ate P-value | AD P-value | Cardiom<br>etabolic<br>Trait<br>P-value | Conditiona<br>l Posterior<br>Probability<br>of<br>Colocalizat<br>ion | Cardiome<br>tabolic<br>GWAS |
| --- | --- | --- | --- | --- | --- | --- | --- | --- | --- | --- | --- | --- |
| DBP | *ADAM10* | rs442495 | 15 | 59022615 | T/C | +/+ | 0.65 | 1.98e-10 | 1.31e-09 | 9.71e-05 | 0.88 | Evangelou et al. 2018 |
| WHRadj<br>BMI | *ADAMTS4* | rs4575098 | 1 | 161155392 | A/G | +/+ | 0.22 | 4.45e-13 | 2.05e-10 | 1.99e-06 | 0.87 | Pulit et al. 2019 |
| DBP | *ACE* | rs4308 | 17 | 61559625 | G/A | +/- | 0.63 | 7.05e-15 | 8.52e-07 | 1.00e-16 | 0.98 | Evangelou et al. 2018 |
| SBP |  | rs4308 | 17 | 61559625 | G/A | +/- | 0.63 | 8.01e-15 | 8.52e-07 | 1.00e-16 | 0.98 | Evangelou et al. 2018 |

To identify potential mechanistic genes at these three loci, we performed single-tissue-eQTL colocalization analysis between the AD signal at each locus using eQTLs identified by GTEx (Materials and Methods). All three pleiotropic signals colocalized with one or more single-tissue eQTL signals (Table S12), and we describe these loci in more detail below.

We detected a pleiotropic signal between AD and DBP at the *ADAM10* locus, discovered as an AD association in Jansen et al. 2019^26^ (Figure S1). Previous single-trait GWAS have identified several other cardiometabolic trait associations near this locus (within a 1MB window around the lead SNP), but our colocalization results suggest that these signals are independent of the AD signal at this locus^35^. Single-tissue eQTL colocalization analysis identified a single eQTL for *MINDY2* in tibial nerve tissue that met our colocalization threshold (Figure S1 and Table S12).

The second pleiotropic signal we detected was at the *ADAMTS4* locus between WHRadjBMI and AD, also discovered in Jansen et al. 2019^26^ AD GWAS (Figure S2). Single-tissue-eQTL colocalization analysis demonstrated that eQTLs for the gene *NDUFS2* across multiple tissues strongly colocalized with this signal (Table S12). An eQTL for the gene *FCER1G* in tibial nerve also met our colocalization threshold (Table S12).

Finally, we detected pleiotropic signals at the *ACE* locus, which is a known blood pressure and AD associated locus, between both DBP and AD and SBP and AD (Figure 2a and Table 1)^29,41^. We noted a direction of effect opposite to the epidemiological correlation for both of these signals, meaning the allele that was associated with reduced risk of AD was associated with increased blood pressure. Our single-tissue-eQTL colocalization showed that both pleiotropic signals had strong evidence of colocalization with eQTLs for *ACE* (Table S12), but also were opposite directions of effect among different tissues (Table 2)^42^.

**Table 2.** Approximate conditional analysis on tissue-specific allelic series in *ACE* eQTLs Approximate conditional analyses performed on the leading single-tissue eQTLs at the *ACE* locus.

| Tissue of *ACE* eQTL | P-value of rs4308<br>conditioned on rs4324 | Effect of rs4308 on the<br>*ACE* eQTL conditioned<br>on rs4324 | P-value of rs4324<br>conditioned on rs4308 | Effect of r rs4324 on the<br>*ACE* eQTL conditioned<br>on rs4308 |
| --- | --- | --- | --- | --- |
| Lung | 0.15 | 0.048 | 2.57e-08 | 0.18 |
| Cerebellum | 5.34e-04 | -0.25 | 0.66 | -0.034 |
| Kidney Cortex | 4.41e-08 | 0.49 | 0.21 | -0.14 |

**Figure 2.**
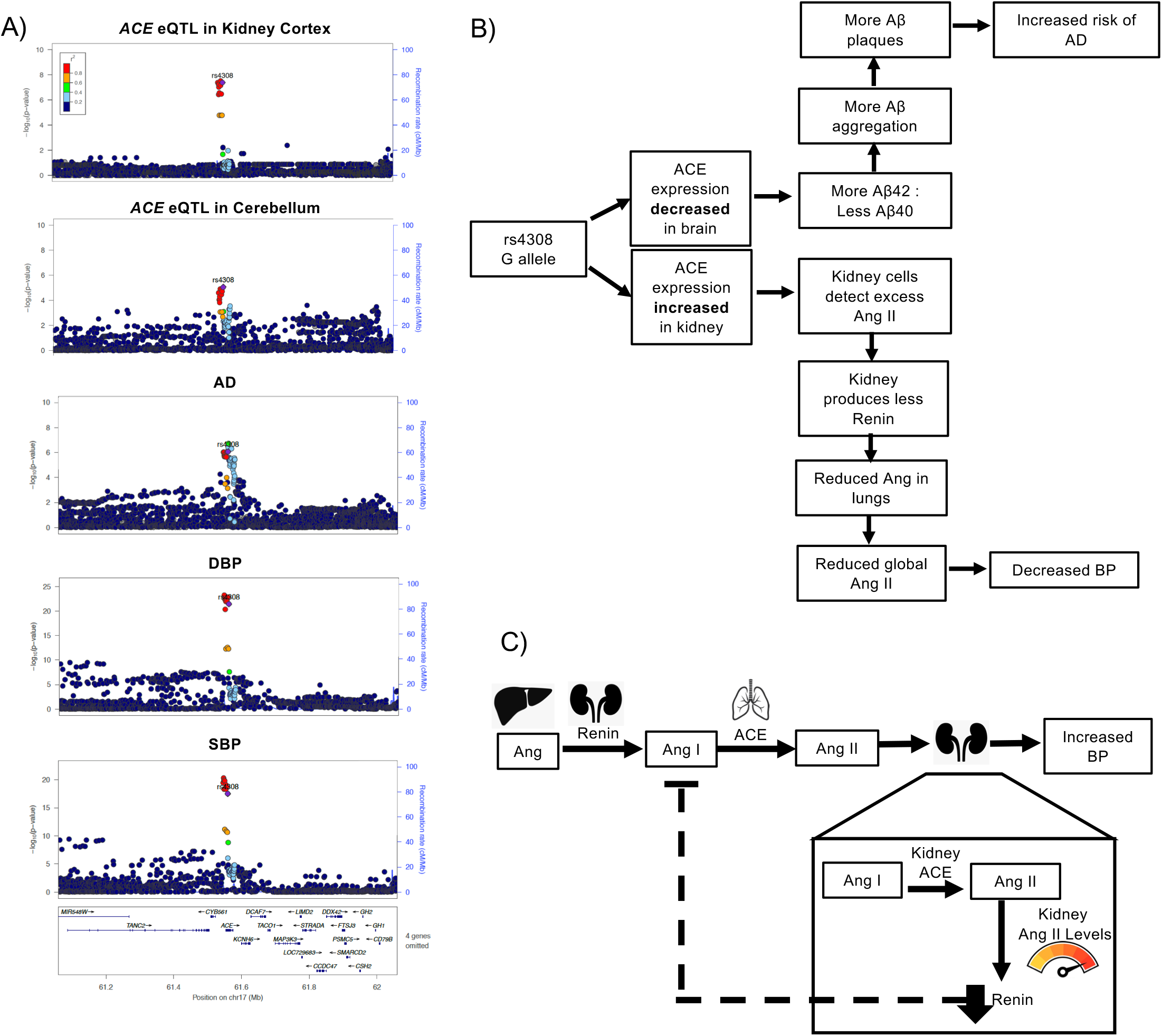
Hypothesized role of the *ACE* in AD risk and hypertension a) Pleiotropic signal between DBP, SBP, and AD at the *ACE* locus, and the eQTL signal for *ACE* in kidney cortex from and cerebellum. b) Flowchart of our hypothesized mechanism as to how tissue-specific expression of *ACE* could mediate the blood pressure (BP) and AD pleiotropic signal at this locus. c) Diagram of a hypothesized mechanism by which increased kidney expression of *ACE* could alter renin expression and thus lead to reduce BP through the feedback loops of the renin-angiotensin system.

The observed complexity of opposite direction effects at this locus motivated us to further investigate the potential of multiple variants associating with traits and/or eQTLs in the region to confound our colocalization analyses. Here, we performed approximate conditional analyses on the pleiotropic signal lead SNP, rs4308, and the lung *ACE* eQTL lead SNP, rs4324, in the single-tissue *ACE* eQTL data for kidney cortex (GTEx v8), lung (GTEx v7), and cerebellum (GTEx v7) (Table 3). The results of this analysis suggested that the *ACE* eQTL in lung was independent of the *ACE* eQTLs in the other tissues. These results also support that the *ACE* eQTLs in kidney and cerebellum share the same causal SNP, which has opposite directions of effect in these tissues (Table 2).

**Table 3.** Tissue-specific *ACE* eQTL colocalization with GWAS trait signals at the *ACE* locus Conditional Posterior Probability of Colocalization, PP4/ (PP3 + PP4) the results of the colocalization analysis between each trait and *ACE* eQTL.

| Tissue of *ACE* eQTL | Conditional probability of colocalization with AD | Conditional probability of colocalization with T2D | Conditional probability of colocalization with DBP | Conditional probability of colocalization with SBP |
| --- | --- | --- | --- | --- |
| Lung | 0.89 | 0.96 | 2.73e-04 | 2.72e-04 |
| Cerebellum | 0.95 | 0.58 | 0.98 | 0.97 |
| Kidney Cortex | 0.97 | 0.22 | 0.99 | 0.99 |

We next assessed which *ACE* eQTLs were most likely to be involved with each of the single-trait signals at this locus, which included the AD, DBP, and SBP signals that we report as pleiotropic as well as a T2D signal that also occurred in this region (Figure S3). We performed colocalization analysis of each of the trait signals with the single-tissue *ACE* eQTLs in kidney cortex, lung, and cerebellum (Table 3). The T2D signal colocalized with the lung *ACE* eQTL, but not with the kidney and cerebellum *ACE* eQTLs. The DBP and SBP signals colocalized with the cerebellum and kidney *ACE* eQTLs, but not the lung *ACE* eQTL. The AD signal colocalized with all three *ACE* eQTLs, but the evidence for colocalization was stronger for the cerebellum and kidney *ACE* eQTLs (Table 3). These results suggest that the blood pressure and AD pleiotropic signals share the same causal SNP that is in high LD with rs4308, and that these associations could be mediated by changes in *ACE* expression in kidney and brain tissue. However, the T2D signal at this locus appears to be independent of the rs4308 signal and could be mediated by changes in *ACE* expression in lung tissue.

## Locus Discovery Analysis Results

We moved to a broad-scale locus discovery effort using bivariate GWAS to detect novel pleiotropic loci that were not previously associated with AD or the eleven cardiometabolic traits of interest (Materials and Methods). After applying a battery of filters to identify the subset of loci with positive evidence of pleiotropy and novelty, we were left with thirteen independent loci (Table S4).

We next performed trait-trait colocalization analysis, and found that three of the thirteen independent loci colocalized. Thus, there was strong evidence of a shared causal SNP between AD and cardiometabolic traits at these loci. Among the thirteen independent loci was a locus with low frequency exonic lead SNP with a bivariate P-value of 7×10^-8^. Due to the lead SNP being a low frequency SNP it had very little LD with other SNPs, which was not conducive to colocalization analyses.

To identify candidate causal genes, we performed single-tissue-eQTL colocalization analysis at the three loci that were conducive to colocalization analysis. We found that all three loci colocalized with one or more single-tissue eQTL signals from GTEx v7 (Table S13).

The first novel pleiotropic signal we detected was between LDL and AD at the *DOC2A* locus (Figure S4). This region has been implicated in other cardiometabolic and neurological traits in previous single-trait GWAS^35,43-45^. The lead SNP, rs11642612, is in LD (1kG EUR r^2^=0.753) with SNPs that are associated with BMI and schizophrenia ^35^. Single-tissue eQTL colocalization found that this pleiotropic signal colocalized with several eQTL signals, but it most strongly colocalized with an eQTL for *DOC2A* in pancreatic tissue (Table S13).

The next pleiotropic signal was between AD and HDL at the *SPPL2A* locus with the lead SNP rs12595082 (Figure S5). This locus was reported as near genome-wide significantly associated with late-onset AD in Kunkle et al. 2019^41^, however this is the first analysis to detect it at genome-wide significance. This locus was also detected (P-value = 7.16×10^-8^) in our AD and DBP bivariate GWAS with the lead SNP rs12440570. Colocalization analysis suggests that the AD, HDL, and DBP association peaks all colocalize with each other (conditional probability of colocalization = 0.81) (Supplemental Materials and Methods: MOLOC for the *SPPL2A* locus)^46^. The single-tissue eQTL analysis showed that this signal colocalized with eQTLs for multiple nearby genes (Table S13).

We detected an opposite direction of effect pleiotropic signal between AD and BFP at the *CCNT2* locus (Figure S6). Several other neurological and cardiometabolic traits have been associated with this locus^35^. The lead SNP, rs10496731, is in LD with SNPs that are associated with Parkinson’s disease (1kG EUR r^2^>0.378), and DBP (1kG EUR r^2^>0.978) from single-trait GWAS^35^. Single-tissue-eQTL colocalization analysis indicated this signal colocalized with eQTLs for *CCNT2* in skin, and *AC016725.4* in testis (Table S13).

The pleiotropic signal we detected at the *DOCK4* locus was between AD and DBP, with rs144867634 as the lead SNP (Figure 3). rs144867634 is a low frequency missense variant that is two bases away from the 3’ splice junction of the eleventh exon of *DOCK4* (Figure 3a). This led us to evaluate whether rs144867634 alters the splicing of *DOCK4*. According to our *in silico* evaluation of rs144867634’s effect on splicing, it is likely that it alters the splicing of *DOCK4*, leading to exon 11 being spliced out of the *DOCK4* transcript (Figure 3) (Supplemental Materials and Methods and Tables S14-S16).

**Figure 3.**
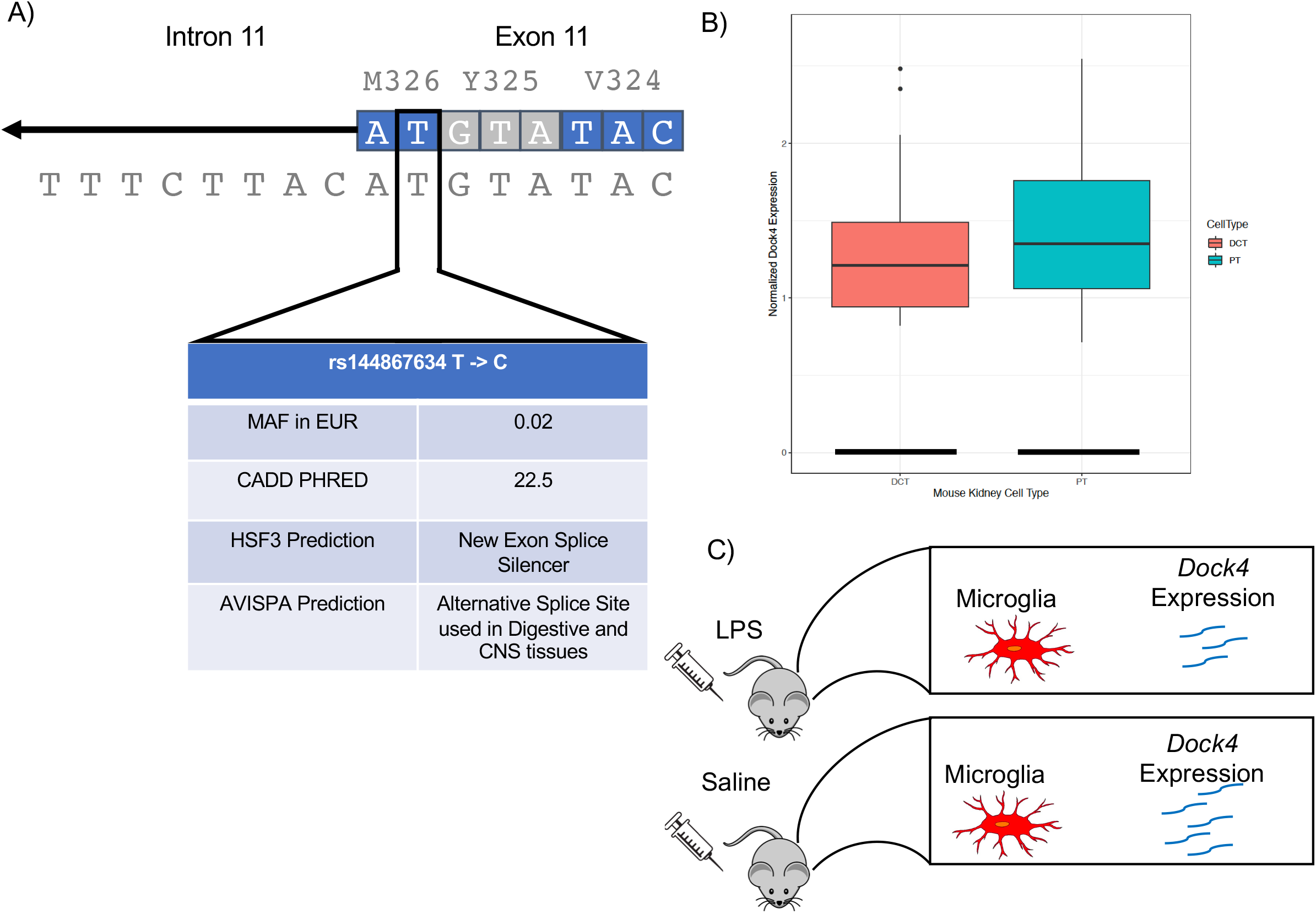
Evidence to support DOCK4 has a role in AD risk and changes in DBP Evidence that supports rs144867634 being the causal variant for the pleiotropic signal at the DOCK4locus. a) In silico evidence that rs144867634 alters DOCK4 splicing. The variant is at the splice junction and is predicted to alter splicing by Human Splice Finder 3 (HSP3) and ASVIPA. b) Single cell mouse kidney data show that Dock4 is expressed by proximal convoluted tubule cells (PT) (128 PT cells of 26,482 assayed have clear evidence of expressing Dock4) and distal convoluted tubule cells (DCT) (27 DCT cells of 8,544 assayed have clear evidence of expressing Dock4) (Park et al. 2018). c) Mouse brain single cell data show that Dock4 expression is reduced in microglia when mice have a neuroinflammatory response induced by endotoxin lipopolysaccharide (LPS) injections (Srinvasan et al.2016).

## Discussion

Here, we demonstrate that a bivariate GWAS method coupled with colocalization analysis enabled the detection of pleiotropic loci between these complex traits and identification of plausible causal genes and potential therapeutic targets. We detected three AD-associated loci with previously unknown pleiotropy for cardiometabolic traits and four loci that were pleiotropic and novel for both AD and the pertinent cardiometabolic trait, all of which we were able to map to one or more potential causal genes.

Our findings support those of previous pleiotropy studies between these traits: that there is a complex genetic relationship between AD and cardiometabolic traits involving both vertical and horizontal pleiotropy^4^. Many of the loci suggest a mechanism where AD and cardiometabolic traits have different causal tissues for the two traits. Further evaluation of the loci we reported could aid in predicting the side effects of medications and for drug repurposing for AD and cardiometabolic diseases.

The potential causal genes we identified through single-tissue-eQTL colocalization analysis support the roles of blood pressure and immune response in both AD and cardiometabolic traits. Three of the pleiotropic loci we report implicate blood pressure mechanisms involved in the pleiotropic relationship at the locus, and four loci had candidate causal genes that have been shown to be involved in immune responses. While these mechanisms make sense given that hypertension and inflammation have both been linked with AD and cardiometabolic diseases, they have not been prevalent in the discussion of pleiotropy between these traits^47-54^.

The pleiotropic signal at the *ACE* locus is a potentially clinically relevant, but complex, finding. *ACE* is an important enzyme in the renin-angiotensin system, and it is the target gene of ACE-inhibitors, a common hypertension medication. We found that the allele associated with increased risk of AD and decreased DBP and SBP was associated with decreased *ACE* expression in brain tissues and most other tissues, but increased *ACE* expression in transverse colon and kidney (Figure 2b and Table S12). These opposite direction of effect single-tissue *ACE* eQTLs appear to colocalize with one another and be independent of a lung *ACE* eQTL nearby (Table 2 and Table 3). However, we cannot exclude the possibility of two causal variants that are both in LD (1kG EUR r^2^ > 0.8) with the lead SNP of the pleiotropic signal, rs4308.

The decrease in blood pressure could be due to the increase in *ACE* expression in the kidney and the negative feedback loop between angiotensin II and renin (Figure 2b,c)^55^. Our hypothesis is that increased expression of *ACE* in the kidney leads to increased levels of angiotensin II in the kidney. These locally increased levels of angiotensin II lead to reduced expression of renin, slowing the entire renin-angiotensin system, and decreasing blood pressure (Figure 2c).

In recent years, the relationship between ACE-inhibitors and AD has been an active field of study and has resulted in two leading hypotheses of how ACE-inhibitors may alter AD risk^48,50,51,56^. Several studies have found that patients on ACE-inhibitors that cross the blood-brain barrier (centrally acting) are at reduced risk of dementia and have improved cognitive ability. Other studies have found evidence that patients taking ACE-inhibitors have decreased cognitive function and increased levels of (β-amyloid (Aβ) protein in their central nervous system; these results were also replicated in mice^51^. This is thought to be due to ACE’s ability to cleave Aβ42 to Aβ40, which is a form of Aβ that is less pathogenic than Aβ42 due to it being less prone to aggregate in the brain^51^. Increases in Aβ42 to Aβ40 ratios have been associated with the *PSEN1* and *PSEN2* mutations in the familial form of AD^57^. Our results support this second hypothesis, that reduced ACE activity in the brain leads to more Aβ42, which in turn leads to more Aβ plaques and an increase in AD risk (Figure 2b). Our findings suggest that further work should be done to evaluate the role of ACE therapeutics for risk of AD.

The BFP and AD pleiotropic signal at the *CCNT2* locus has a particularly compelling potential mechanism. Single-tissue-eQTL colocalization analysis detected colocalization between the bivariate signal and an eQTL for *CCNT2* in skin tissue (Figure S6 and Table 4). The gene *CCNT2* is a strong candidate for being involved with both the BFP and the AD association. *CCNT2* has been shown to be important in adipose biology^58^. Human *CCNT2* knockout adipocytes have altered adipogenesis gene expression and decreased secretion of the hunger inhibiting hormone leptin, which is consistent with increased BFP^58^. CCNT2 has also been shown to be used by herpes simplex virus 1 (HSV-1) when it transcribes its genome^59^. This is a plausible link to AD due to the hypothesis that HSV-1 can trigger amyloid plaques^60-62^.

**Table 4.** Locus discovery analysis pleiotropic loci Chr, chromosome of the SNP; Direction of effect first position is the direction of effect of the effect allele on AD and the cardiometabolic trait; Effect Allele Frequency from the Jansen et al. 2019 allele frequency; Conditional Posterior Probability of Colocalization, PP4/ (PP3 + PP4) the results of the trait-trait colocalization analysis.

| Cardiom etabolic Trait | Locus Name | Lead SNP | Chr | Position GRCh37 | Effect Allele/ Other Allele | Directi on of Effect AD/CM | Effect Allele Frequency | Bivari ate P-value | AD P-value | Cardiom etabolic Trait P-value | Conditional Posterior Probability of Colocalizati on | Cardiom etabolic GWAS |
| --- | --- | --- | --- | --- | --- | --- | --- | --- | --- | --- | --- | --- |
| LDL | *DOC2A* | rs11642612 | 16 | 30030195 | A/C | +/+ | 0.62 | 2.19e-08 | 5.32e-06 | 2.68e-06 | 0.98 | Klarin et al. 2018 |
| HDL | *SPPL2A* | rs12595082 | 15 | 51007729 | C/T | +/+ | 0.81 | 3.27e-08 | 1.66e-06 | 1.09e-05 | 0.95 | Klarin et al. 2018 |
| BFP | *CCNT2* | rs10496731 | 2 | 135597628 | G/T | +/- | 0.48 | 1.74e-08 | 1.72e-05 | 2.67e-05 | 0.94 | Lu et al. 2016 |
| DBP | *DOCK4* | rs144867634 | 7 | 111580166 | T/C | +/+ | 0.98 | 8.33e-08 | 5.36e-05 | 1.089e-07 | NA | Evangelou et al. 2018 |

Finally, our results suggest that *DOCK4* is the putative causal gene for the pleiotropic signal between DBP and AD at the *DOCK4* locus, since the lead SNP is a low frequency exonic variant in *DOCK4* that is predicted to lead to exon 11 of *DOCK4* being spliced out of the *DOCK4* transcript (Figure 3a). For these reasons, and the fact that the rare allele is associated with lower risk of AD and reduced DBP, *DOCK4* is our strongest candidate for a novel therapeutic target. The human genetics data observed here is consistent with the simple hypothesis that reduced efficacy of *DOCK4 in vivo* could treat both hypertension and AD. There is already evidence that *DOCK4* could be involved with AD and DBP. Previous genetic studies have shown that *DOCK4* variants are associated with multiple neurological phenotypes, and *DOCK2*, the other member of *DOCK4’s* protein subfamily, expression is increased in the microglia of patient’s with AD^63-67^. It has also been shown that Dock4 expression in mouse microglia is altered when mice are given an endotoxin lipopolysaccharide (LPS) injection to induce a neuroinflammatory response (Figure 8c)^68^. *DOCK4* could also affect DBP through changes in kidney function. *DOCK4* is expressed in kidney in GTEx v8, and *Dock4* is expressed in mouse kidney proximal tubule cells and distal convoluted tubule cells. These cells are responsible for reabsorption of salts, sugars, and amino acids in the nephron of the kidney, and thus altering their function could change blood volume (Figure 3b)^40,69^.

There are several limitations of our study. The Jansen et al. 2019^26^ AD GWAS and many of the cardiometabolic trait GWAS we used included individuals from the UK Biobank dataset. This sample overlap will increase the estimated covariance between our traits making the resulting bivariate P-value more conservative for a locus that has the same direction of effect as the phenotypic correlation and less conservative when a locus has an opposite direction of effect. The overlapping samples may also inflate our posterior probability of colocalization between these traits. A phenotypic limitation is that the Jansen et al. 2019^26^ data includes some Proxy-AD patients.

## Conclusion

We have shown that bivariate GWAS paired with colocalization analysis can be an effective way to detect pleiotropic loci between complex traits and generate hypotheses as to why these loci are pleiotropic. We detected seven loci that have evidence of being pleiotropic between AD and a cardiometabolic trait, and we were able to identify candidate causal genes for all of these loci. Two loci seem to stand out in their potential to improve our ability to prevent and treat AD. The first is the *ACE* locus, which provides more evidence to support a potential link between AD risk and ACE-inhibitors. The other is the *DOCK4* locus which is our most promising candidate for a novel therapeutic target. Our results may aid in resolving the etiology of AD, and help identify new therapeutic targets for this disease. AD is a complex disease, and we expect that applying this method to other traits that have been associated with AD, such as educational attainment and immune traits, should also lead to novel pleiotropic loci, new candidate causal genes, and a better understand of AD^70-73^.

## Data Availability

GWAS summary statistics data used in the paper are available at: AD data PMID:30617256, BFP data PMID:26833246, BMI data PMID: 30124842, CHD data PMID: 29212778, DBP data PMID: 30224653, HDL data PMID: 30275531, LDL data PMID: 30275531, SBP data PMID: 30224653, TC data PMID: 30275531, TG data PMID: 30275531, T2D data PMID: 30297969, WHRadjBMI data PMID: 30239722
Access to the MVP lipids data can be obtained from dbGAP (phs001672.v4.p1, pha004828.1, pha004831.1, pha004837.1, pha004834.1) and GLGC European ancestry only data can be obtained at: http://csg.sph.umich.edu/willer/public/lipids/ or http://lipidgenetics.org/.

## Acknowledgements

We want to thank Theodore G. Drivas, Tiffany R. Bellomo, Jason E. Miller, and Pankhuri Singhal for sharing their medical and phenotypic knowledge. This research is based on data from the Million Veteran Program, Office of Research and Development, Veterans Health Administration, and was supported by the Department of Veterans Affairs Office of R&D award I01-BX003362 (to P.S.T. and K.M.C) and IK2-CX001780 (to S.M.D). This publication does not represent the views of the Department of Veteran Affairs or the United States Government. This work was supported by the American Heart Association (20PRE35120109 to W.P.B.), the National Institutes of Health (DK101478 to B.F.V. and P50GM115318-01 to M.D.R.), and a Linda Pechenik Montague Investigator Award (to B.F.V.).

## Web Resources

GWAS Catalog, https://www.ebi.ac.uk/gwas/docs/file-downloads

LocusZoom, http://locuszoom.org/genform.php

LDlink, https://ldlink.nci.nih.gov/?tab=ldassoc

MyVariant.info, https://myvariant.info/

Combined Annotation Dependent Depletion (CADD), https://cadd.gs.washington.edu/score

Human Splice Finder 3, http://www.umd.be/HSF3

## Data and Code Availability

The bivariate GWAS code generated during this study are available at AD_and_Cardiometabolic_Trait_Bivariate_Scans

https://github.com/wpbone06/AD_and_Cardiometabolic_Trait_Bivariate_Scans.

The eQTL colocalization code generated during this study are available at GTEx_v7_eQTL_colocalizer https://github.com/wpbone06/GTEx_v7_eQTL_colocalizer.

GWAS summary statistics data used in the paper are available at: AD data PMID:30617256, BFP data PMID:26833246, BMI data PMID: 30124842, CHD data PMID: 29212778, DBP data PMID: 30224653, HDL data PMID: 30275531, LDL data PMID: 30275531, SBP data PMID: 30224653, TC data PMID: 30275531, TG data PMID: 30275531, T2D data PMID: 30297969, WHRadjBMI data PMID: 30239722

Access to the MVP lipids data can be obtained from dbGAP (phs001672.v4.p1, pha004828.1, pha004831.1, pha004837.1, pha004834.1) and GLGC European ancestry only data can be obtained at: http://csg.sph.umich.edu/willer/public/lipids/ or http://lipidgenetics.org/.

## Supplemental Data Description

6 Supplemental Figures in the Supplemental Data PDF

16 Supplemental Tables: 5 in the Supplemental Data PDF (Tables S1-S8, S10-S12, S14) and 2 in Excel sheets (Tables S9 and S13)

## Notes

### Competing Interest Statement

The authors have declared no competing interest.

### Author Declarations

University of Pennsylvania

